# Social heterogeneity and the COVID-19 lockdown in a multi-group SEIR model

**DOI:** 10.1101/2020.05.15.20103010

**Authors:** Jean Dolbeault, Gabriel Turinici

## Abstract

The goal of the lockdown is to mitigate and if possible prevent the spread of an epidemic. It consists in reducing social interactions. This is taken into account by the introduction of a factor of reduction of social interactions q, and by decreasing the transmission coefficient of the disease accordingly. Evaluating *q* is a difficult question and one can ask if it makes sense to compute an average coefficient *q* for a given population, in order to make predictions on the basic reproduction rate ℛ_0_, the dynamics of the epidemic or the fraction of the population that will have been infected by the end of the epidemic. On a very simple example, we show that the computation of ℛ_0_ in a heterogeneous population is not reduced to the computation of an average *q* but rather to the direct computation of an average coefficient ℛ_0_. Even more interesting is the fact that, in a range of data compatible with the Covid-19 outbreak, the size of the epidemic is deeply modified by social heterogeneity, as is the height of the epidemic peak, while the date at which it is reached mainly depends on the average ℛ_0_ coefficient. This paper illustrates more technical results that can be found in [4], with new numerical computations. It is intended to draw the attention on the role of heterogeneities in a population in a very simple case, which might be difficult to apprehend in more realistic but also more complex models.

## The model

We consider a compartmental model based on the SEIR equations (for *Susceptible, Exposed, Infected, Recovered*) with *n* categories of susceptible individuals and a total number of individuals *N* given by

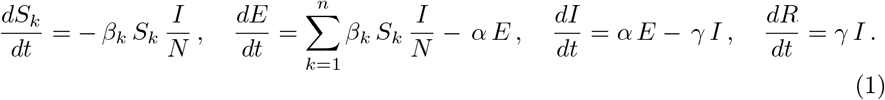

We consider this system as a function of *t* ≥ 0 in terms of the initial data corresponding to the values of *S, E, I* and *R* at *t* = 0. The *average incubation period* is 1*/α*, the parameter *β*_*k*_ is the product of the average number of contacts per person and per unit time by the probability of disease transmission in a contact between a susceptible individual in the group *k* and any infectious individual, *γ* is a *transition rate* so that 1*/γ* measures the duration of the infection of an individual (or actually how long he is infectious and able to contaminate other people before being isolated), and *N* is the total population size.

An individual in the group *k* is characterized by his *transmission rate β*_*k*_. In simple SEIR models of lockdown, a single group is considered (*n* = 1) and it has been proposed for instance in [7] to introduce a *factor of reduction of social interactions q* so that the effect of the lockdown is to reduce the transmission rate from *β*_1_ before lockdown to *β*_1_*/q* after lockdown. Our goal is to study what happens if there are, after lockdown, *n* ≥ 2 groups with different *factors of reduction of social interactions q*_*k*_, so that

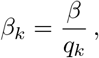

where *β* is a fixed, given parameter. We shall speak of *social groups* and *social heterogeneity* because each group has its own transmission rate *β*_*k*_. This rate does not interfere with the dynamics of the disease once the corresponding individual is infected. In a period of lockdown, the reduction factor is not the same for a health worker, a supermarket cashier or an employee working from home by internet. Actually *β*_*k*_ can also be used to take into account other characteristics of the population, like age groups, which play a role in the risk of becoming infected. Here we assume that *q*_*k*_ determines the initial group of an individual and does not vary over time. The other coefficients *α, β* and *γ* of System (1) are also assumed to be independent of *t*. A last point of terminology concerns the notion of individuals. As we use a compartmental model, we should speak only of the proportion of individuals in a compartment. Since the total number of individuals *N* is taken large, this proportion can be considered as a continuous variable subject to ordinary differential equations, which is precisely what we do in (1).

### Reduction of social interactions and basic reproduction ratio

Let us consider the initial probability distribution among the groups such that, at *t* = 0,

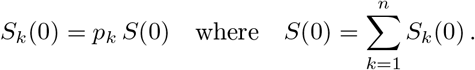

Notice that *p*_*k*_ is a parameter used for the description of the initial datum only. In a *disease free equilibrium* corresponding to *S* = *N*, it would be very natural to consider an average *factor of reduction of social interactions*

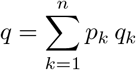

and this is actually what is done implicitly when a single compartment of susceptible individuals is considered. In that case, the *basic reproduction ratio* computed for instance by the *next generation matrix method* (see [3] and references therein) is *β/*(*q γ*). However, if we apply the next generation matrix method to (1), we found in [4, Theorem 4.2 (1)] that the *basic reproduction ratio* is

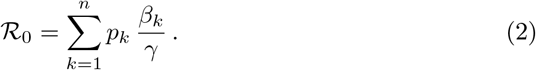

Although this is a relatively elementary property, there are already important consequences, for instance in the case 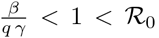. While a model with a single compartment and an average factor of reduction of social interactions predicts the extinction of the epidemic, a small group with high transmission factor *β*_*k*_, even if numerically not very large (that is, with *p*_*k*_ small), can spread the disease and trigger an outbreak. This is one of the possible interpretations of the data, which were suggesting a pattern corresponding to a basic reproduction ratio larger than 1 in the initial stage of the lockdown in France rather than an expected exponential decay of the number of cases.

### Limitations and choice of the numerical parameters

System (1) is an extremely simplified model, in which *Undetected* or asymptomatic cases are not taken into account, although this seems an important issue in the Covid-19 pandemic. It is also a model for short term (say of the order of three months) so that the evolution of the structure as well as natural birth and death rates are not taken into account. If Covid-19 becomes endemic or last longer, it will not be possible to keep ignoring such issues anymore. Feedback mechanisms and changes in the values of the parameters due to the evolution of the disease or the changes in social habits are certainly going to play a role. More important is the fact that the parameters are fitted on the basis of official data during the very early stage of the disease.

System (1) is homogeneous so that we can simply consider the *fractions*

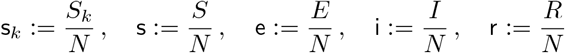

of the *Susceptible, Exposed, Infected* and *Recovered* individuals among the whole population as a function of the time *t* ≥ 0. Numerical values are taken from [2]: for the initial data, at *t* = 0, we consider a perturbation of the *disease free equilibrium* given by s_*k*_(0) = *p*_*k*_ s(0) and

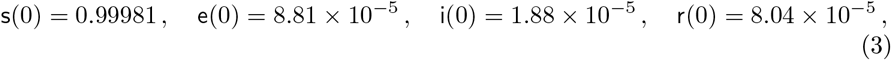

based on the data of March 15, 2020 in France from [8], and choose in (1)

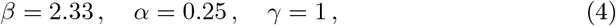

which gives us a basic reproduction ratio of 2.33 (in the case of a single group). The reader interested in further details and comparisons is invited to refer to [4], where the choice of the numerical parameters is discussed in greater details. This set of initial data and parameters is however not a key issue for our discussion. Our contribution is focused on understanding the theoretical implications of social heterogeneity and remains valid for other sets of numerical choices. Numerical choices (3)-(4) are given only for an illustrative purpose.

In [2], N. Bacaër is able to fit the curve of the number of infected individuals with a single group and *q*_1_ = 1.7, at the beginning of the lockdown. In [4], it is shown that with two groups, *p*_1_ = 98%, *p*_2_ = 2%, *q*_1_ = 2.35 and *q*_2_ = 0.117, we can recover the same reproduction ratio of 1.37 as in [2]. In order to fix ideas, we take

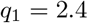

which corresponds to a reduction of the transmission rate of 1*/q*_1_ 42% in the group *k* = 1. We emphasize that we have no empirical basis to determine this parameter. The idea is to pick for *q*_1_ a value larger than 2.33 so that, in presence of a single group, the epidemic would rapidly extinguish. However, a second group with a moderate or high transmission coefficient *β*_2_ is enough to produce an outbreak corresponding to a global ℛ_0_ > 1 and we study how various features of the epidemic curve depend on *β*_2_. See Fig. 1.

**Figure 1.**
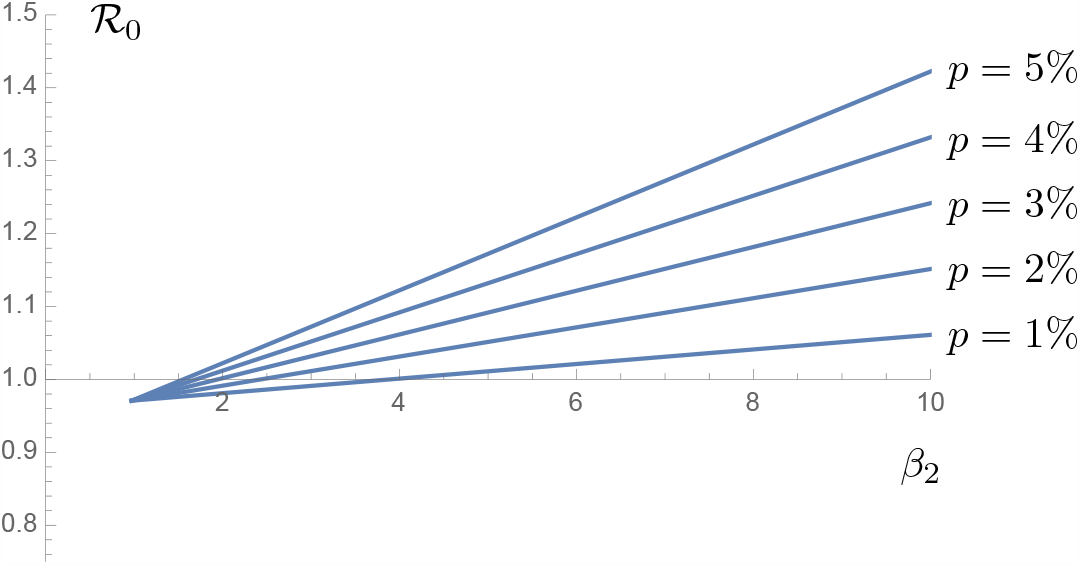
Numerical values of ℛ_0_: we take *p* small but assume that the individuals in the group *k* = 2 may have a moderate or high transmission coefficient *β*_2_.

From now on, we shall assume in all our numerical results that *n* = 2 (two groups) and vary *p* = *p*_2_ (so that *p*_1_ = 1 − *p*). Consistently with the idea that the initial stage of the outbreak is independent of the groups, we take as initial data s_1_(0) = (1 − *p*) s(0) and s_2_(0) = *p* s(0). We keep *p* small and take it in the range 1 to 5% in our examples. It is not difficult to understand that the destabilization due to a small fraction *p* of the population (the *k* = 2 group) in the large group (the *k* = 1 group) in which the epidemic would be under control if it were isolated, is all the stronger when the basic reproduction ratio (inside the *k* = 1 group, considered as isolated) is close to 1: this is the reason why we arbitrarily choose *q*_1_ = 2.4. The case of *q*_2_ small (eventually smaller than 1), corresponding to individuals in the group *k* = 2 with high transmission rates, is of particular interest.

Our model is overly simplified. More realistic models should describe social interactions with far more details, address not only lockdown but also mask wearing and disinfection measures, social-distancing, self-quarantine, various types of stay-at-home orders and curfews, *etc*., and also take into account the evolution of the parameters induced by behavioural changes and economical constraints as studied for instance in [6].

### The epidemic size

The (final) *epidemic size ζ* is defined as the fraction of individuals that are affected by the epidemic for large times, here s(0) − s^⋆^, where s^⋆^ is the fraction which is not infected at the end of the epidemic. Under the condition that ℛ_0_ > 1 and 1 − s(0) is small, we find that r^⋆^ = 1 − s^⋆^ is of the order of the solution *r* of

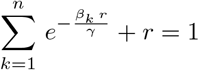

(see in [4, Section 3.2 and Theorem 4.2 (1)]). It is also proved in [4, Theorem 4.2 (3)] that *ζ* is decreased when *n* is increased from *n* = 1 to an arbitrary *n* ≥ 2 (for our numerical examples, we take *n* = 2). See Figs. 2 and 3. In Fig. 3, ℛ_0_ = 1.37 is achieved in a single group with *q* ≈ 1.70 as in [2], which gives an epidemic size *ζ* ≈ 49%. The same value ℛ_0_ = 1.37, which is obtained in two groups with *p* = 1%, *q*_1_ = 2.4 (and *q*_2_ ≈ 0.06), gives an epidemic size *ζ* ≈ 12%.

**Figure 2.**
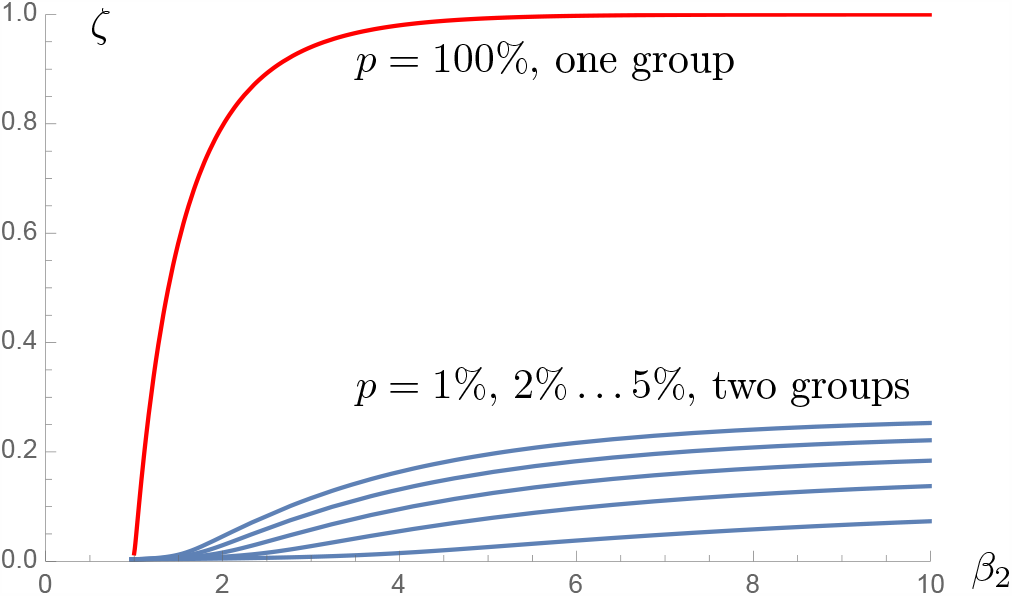
Epidemic size *ζ* as a function of *β*_2_ for *p* = 0.01, 0.02,… 0.05 (blue; *n* = 2) and *p* = 1 (red, *n* = 1). This red curve corresponds to an outbreak with a basic reproduction ratio *β*_2_*/γ*. This is however not so instructive as the *k* = 2 group is a small proportion of the population, but with high transmission rate.

**Figure 3.**
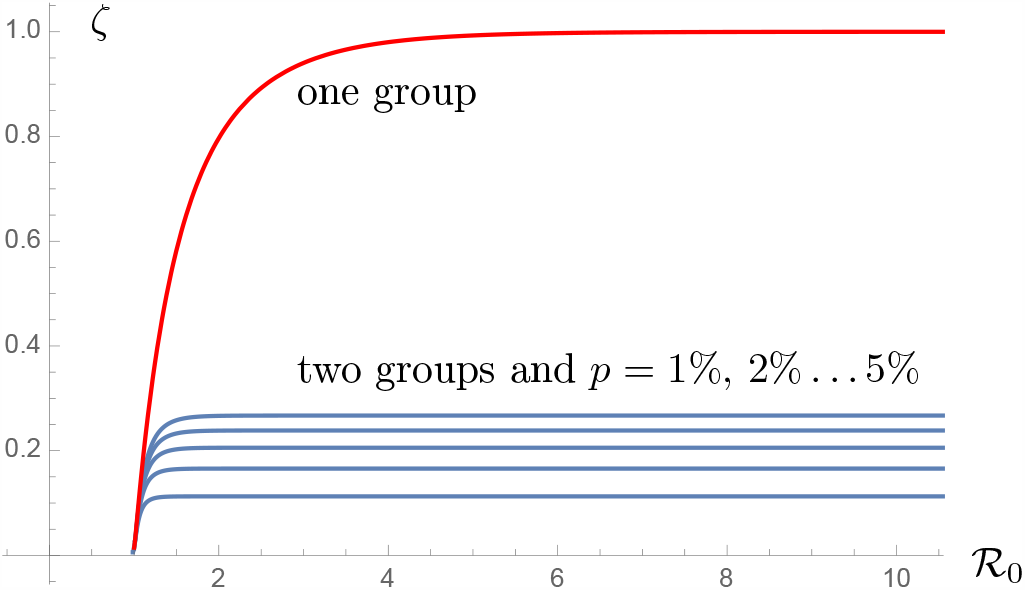
Epidemic size *ζ* as a function of ℛ_0_ for *p* = 0.01, 0.02,. 0.05 (blue) and *p* = 1 (red), which corresponds to the case of a single group. With the same ℛ_0_ = 1.37, we see that *ζ* is drastically decreased.

### The epidemic peak

Not only the asymptotics of the epidemic are changed when replacing the model with *n* = 1 by a model with *n* = 2, but also the dynamical behavior of the solutions. The height of the epidemic peak (defined as the maximum of infected e + i as function of time) is decreased when *n* is increased form *n* = 1 to *n* ≥ 2: see [4] for a proof. The height and date of the epidemic peak strongly depend on *p*. See Figs. 4, 5 and 6. In Fig. 5, for the same value of ℛ_0_ ≈ 1.5, the curve *t*↦ e(*t*) + i(*t*) is represented in red for a single group (this corresponds here to a factor of reduction of social interactions *q* ≈ 1.55) and in blue in the case of two groups (with parameters *q*_1_ = 2.4, *q*_2_ = 0.2 and *p* = 0.05).

**Figure 4.**
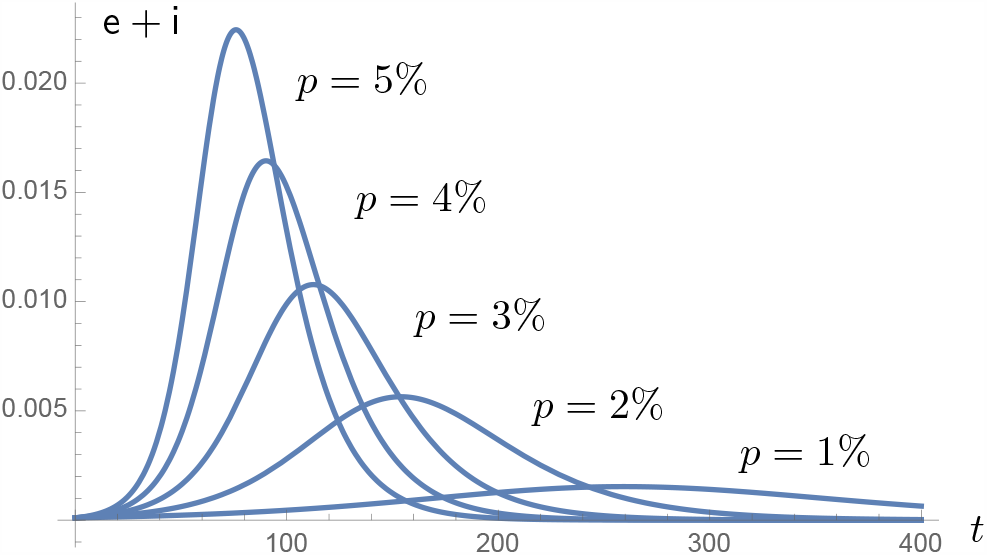
Fraction of exposed and infected individuals e + i as a function of time (in days) for *p* = 0.01, 0.02,… 0.05, with *β*_2_ = 11.65 corresponding to *q*_2_ = 0.2.

**Figure 5.**
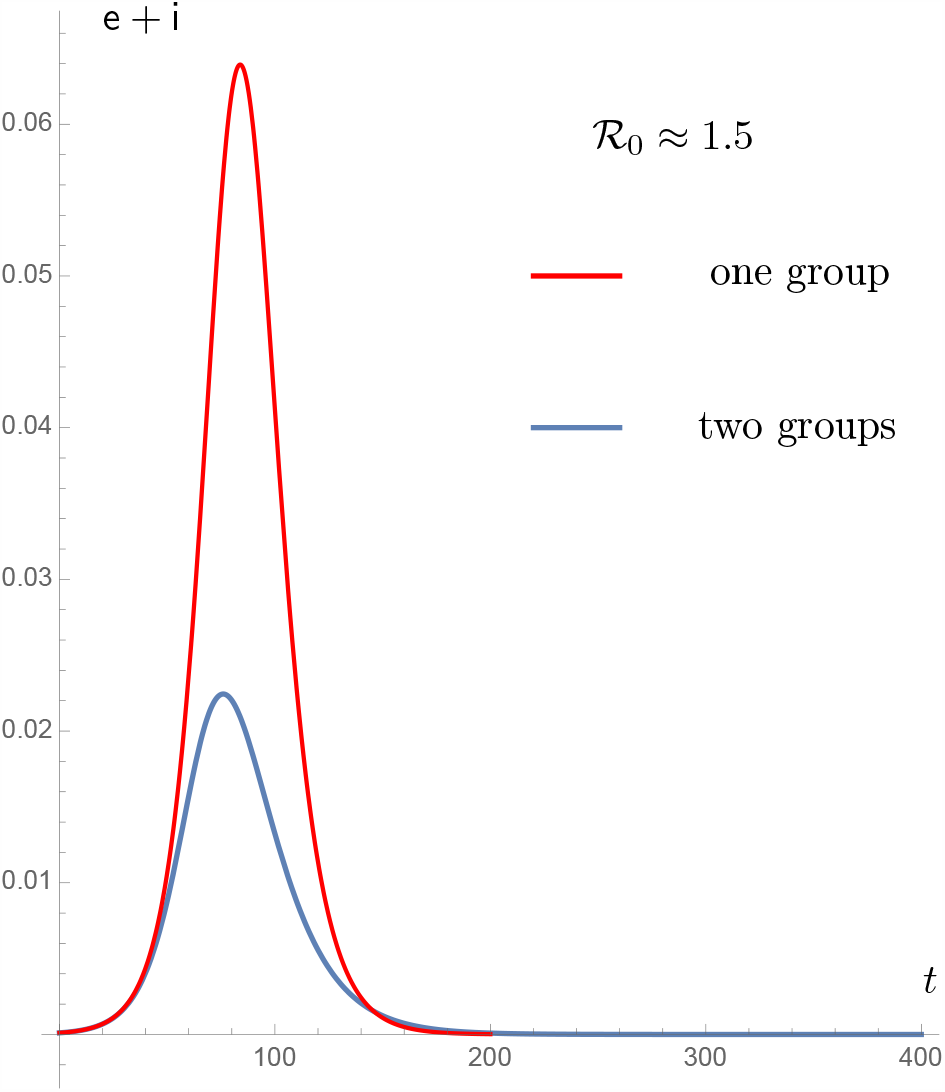
A striking effect of the heterogeneity is the flattening of the curve shown here for ℛ_0_ ≈ 1.5.

**Figure 6.**
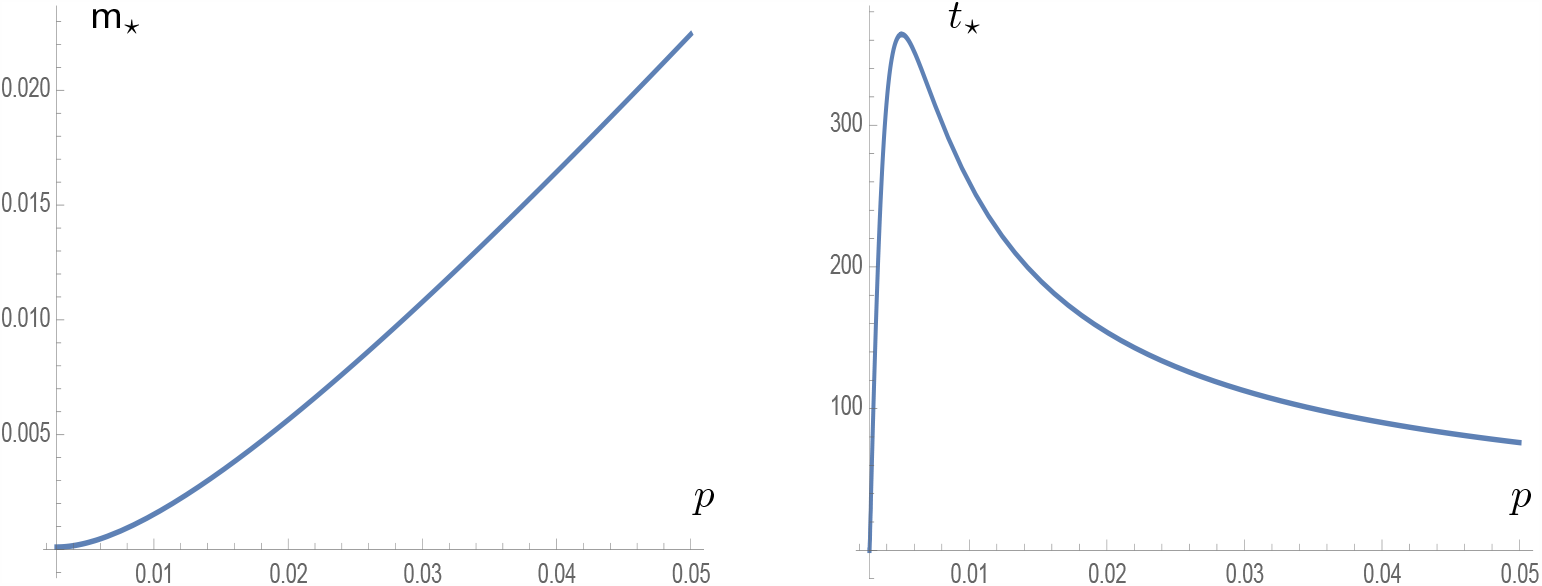
Size (left) and date (right) of the epidemic peak as function of *p* in the range 0.27% to 5%. With our choice of the parameters (*q*_1_ = 2.4 and *q*_2_ = 0.2), there is no outbreak if *p ≤* 0.27%, because ℛ_0_ *≤* 1 in that case.

Now, let us describe how the epidemic peak depends on the parameters in our two groups example. For a given value of *β*_2_, larger values of *p* mean a larger ℛ_0_, with a linear dependence given by (2), a larger epidemic size (see Fig. 2), but also a larger epidemic peak (see Fig. 6). Let us denote by *t*_⋆_ the date of the peak and let m_⋆_ := e(*t*_⋆_) + i(*t*_⋆_) = max_*t*≥0_ e(*t*) + i(*t*). We observe that *p ↦*m_⋆_(*p*) is increasing with *p* and *t*_⋆_ is non-increasing with ℛ_0_ for sufficiently large values of ℛ_0_.

It is also interesting to make comparisons with the same value of ℛ_0_. In our model case for *n* = 2, using (2) and the definition of *β*_*k*_, we can for instance choose *q*_2_ as a function of *p* to achieve a fixed value of ℛ_0_ and get

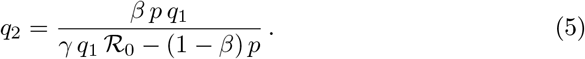

See Fig. 7 for some qualitative results on the dependence of m_⋆_ and *t*_⋆_ in ℛ_0_, for various values of *p*. Although not linear, the size of the epidemic peak is a clearly increasing function of *p* in the outbreak regime. It is an empirical but remarkable fact that, for our set of data, the date of the epidemic peak almost does not depend on *p*.

**Figure 7.**
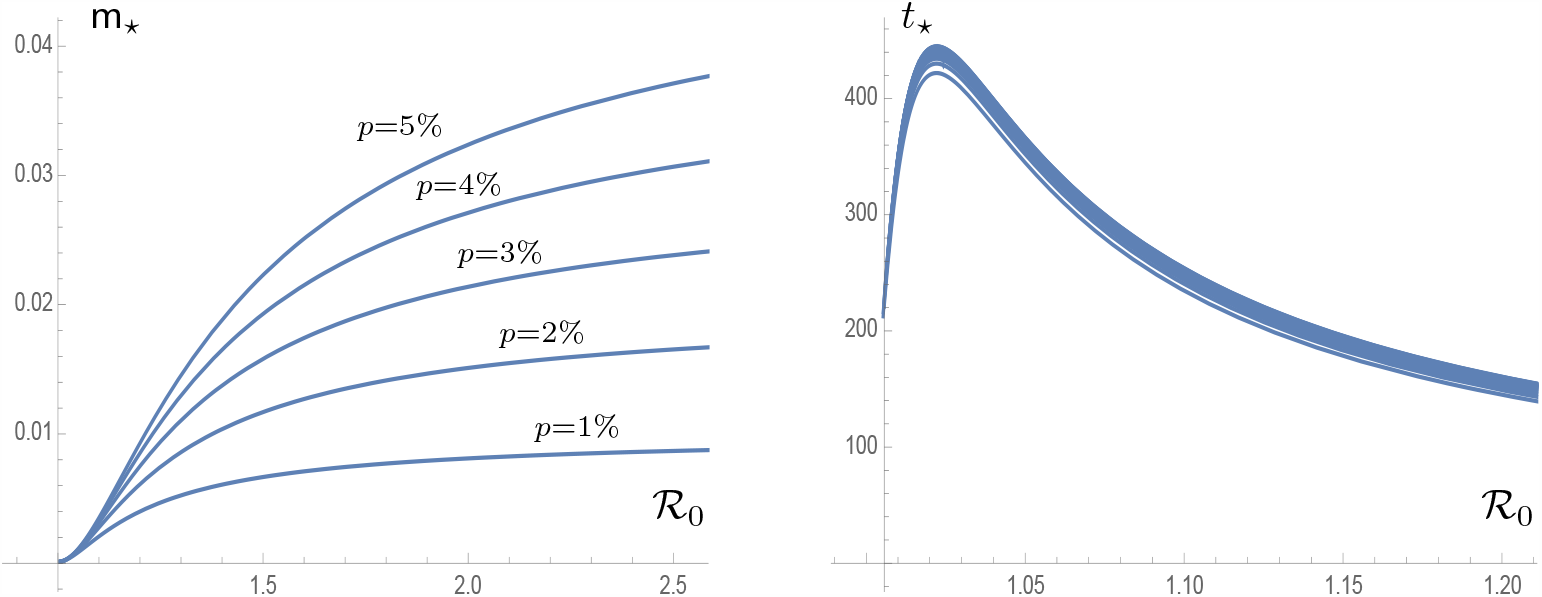
Left: size of the epidemic peak as a function of ℛ_0_. Right: date of the epidemic peak as a function of ℛ_0_. Plots correspond to *p* = 0.01, 0.02,… 0.05.

**Figure 8.**
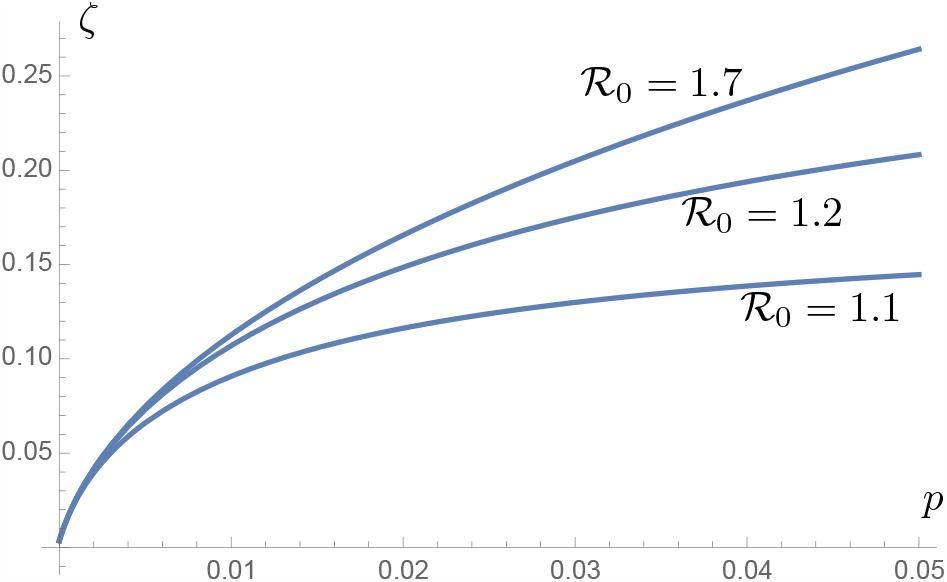
The epidemic size *ζ* as a function of *p* ranging from 0 to 0.05, where *q*_2_ is adjusted according to (5) so that ℛ_0_ is given and equal to 1.1, 1.2 and 1.7.

**Figure 9.**
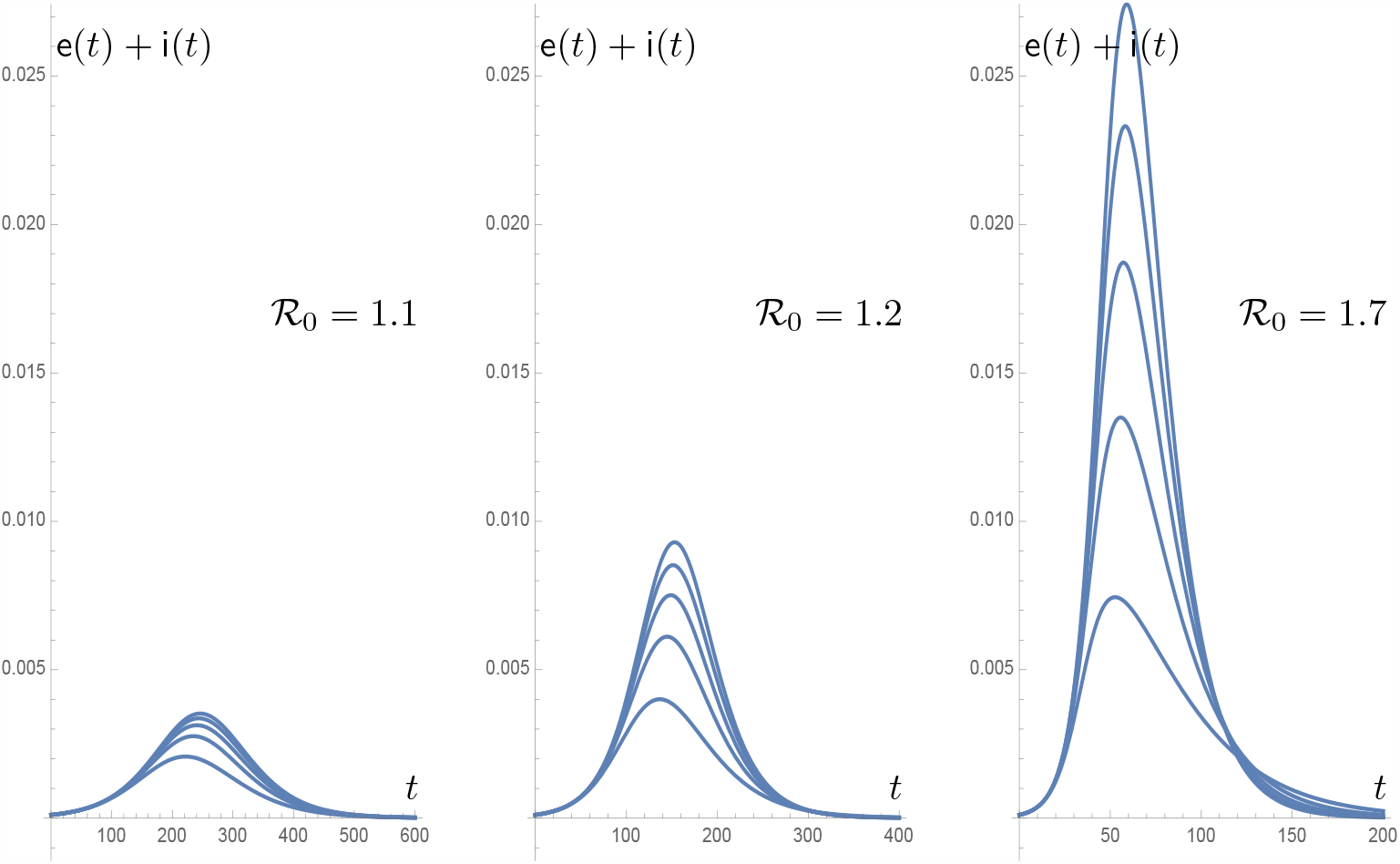
The epidemic curve and the peak strongly depends on ℛ_0_. Notice that the scale on the vertical axis is the same in the three cases corresponding to ℛ_0_ = 1.1, 1.2 and 1.7, while the time scale is respectively (0, 200), (0, 400), and (0, 600). On each plot, the curves correspond to *p* = 0.01, 0.02,… 0.05.

### The ℛ_0_ is not the message

As seen in Fig. 7, ℛ_0_ does not retain all relevant information and for a given *q*_1_ and ℛ_0_, one may still vary either *p* or *q*_2_ and eliminate the other one using (2). Here we shall rely on (5) for a few more plots which show some additional qualitative features of the epidemic size and peak.

## Discussion

The goal of a lockdown is to reduce the basic reproduction ratio of the epidemic to a value less than 1 and drive the disease to extinction, or at least decrease it in such a way that “the curve is flattened”. The efficiency of the lockdown is achieved by a reduction of social interactions, which is measured (in average) by a factor *q*. In this paper we question whether such an average factor makes sense or not. Beyond the difficult issue of giving realistic values to *q*, we study some consequences of social heterogeneities when the population is divided into groups for which *q* takes different values.

The basic reproduction ratio ℛ_0_ has been computed in [4] and is given in (2) as an average of the basic reproduction ratios for each group (or, equivalently, using an average transmission rate) and not as a global ratio based on an average *q* factor,as it it is implicitly done in many papers in the literature. A small group with a high transmission rate eventually triggers an outbreak even if the basic reproduction ratio of the majority is below 1. This papers focuses on some quantitative consequences for well chosen numerical examples.

The qualitative properties of the curves in presence of heterogenous groups are not the same as when considering a single group with *the same* (averaged) basic reproduction ratio. The dynamics of the outbreak and its properties, for instance the height of the epidemic peak er the final epidemic size *ζ*, are also changed. A model with only one group and fitting the observed data in the initial phase of the outbreak will be more pessimistic concerning the epidemic outcomes than a heterogeneous model. This is even more true after lockdown when social distancing measures have been enforced, the lockdown being by its nature a source of heterogeneity as some individuals are exempted. In terms of public health, this also underlines the importance of targeting prevention measures on individuals with a high level of social interactions. After this study has been completed, important epidemiologic data have been published in [5] which point in the same direction.

## Data Availability

No private data has been used in this work.

## Acknowledgment

This work has been partially supported by the Project EFI (ANR-17-CE40-0030) of the French National Research Agency (ANR). The authors thank the [MODCOV19](https://insmidirect.math.cnrs.fr/spip.php?article3647) platform [1] for support. The authors thank a referee for very useful comments and especially for pointing an inconsistency due a very inconvenient typo error.

© 2020 by the authors. This paper may be reproduced, in its entirety, for non-commercial purposes.

## Author’s Statement

Authors state no conflict of interest.

